# Seroresponse to third doses of SARS-CoV-2 vaccine among patients receiving maintenance dialysis

**DOI:** 10.1101/2022.01.03.21268549

**Authors:** Caroline M. Hsu, Eduardo K. Lacson, Harold J. Manley, Gideon N. Aweh, Dana Miskulin, Doug Johnson, Daniel E. Weiner

**Affiliations:** Tufts Medical Center, Boston, MA; Dialysis Clinic Inc., Nashville, TN

**Keywords:** Dialysis, end-stage kidney disease, COVID-19, vaccination, hemodialysis, peritoneal dialysis

## Abstract

Given the increased morbidity and mortality from COVID-19 among patients undergoing maintenance dialysis, we examined the seroresponse to an additional dose of SARS-CoV-2 mRNA vaccine, using a hypothesized protective anti-spike protein IgG antibody threshold of 7 based on prior work. Among 395 patients, 384 (97%) had seroresponse ≥7 following administration of the third dose. Among those with suboptimal (peak <7) and minimal (peak <1) seroresponse to an initial mRNA vaccine regimen, the rate of seroresponse ≥7 following a third dose was 97% (36 of 37) and 64% (14 of 22), respectively. Given ongoing high rates of COVID-19 and rapidly waning immunity after an initial vaccine series, we recommend a third mRNA vaccine dose be standard for all patients receiving maintenance dialysis.

## Introduction

Beginning August 13, 2021, third doses of SARS-CoV-2 mRNA vaccine were recommended for patients with immunocompromise,^1^ possibly but not definitively including patients undergoing dialysis,^2^ who have increased risk for poor outcomes from COVID-19.^3^ In this study, we examined the seroresponse to an additional dose of vaccine in this vulnerable population.

## Methods

Dialysis Clinic, Inc. (DCI) is a national not-for-profit provider caring for approximately 15,000 patients at 260 outpatient dialysis clinics in 29 states. As previously described, beginning in January 2021, DCI physicians had the option of activating a SARS-CoV-2 vaccine protocol, in which anti-spike immunoglobulin G (anti-spike IgG) antibodies were drawn monthly with routine labwork.^4^ This study includes all adult patients who received an additional SARS-CoV-2 vaccine dose after completion of an initial series, and who had at least one anti-spike IgG level checked both between the initial series and the additional dose, and at least 14 days after the additional dose. Demographic and clinical data, vaccination dates, history of COVID-19 diagnosis, and anti-spike IgG titer results were obtained from the DCI electronic health record.

For each patient, three titer levels of interest were identified: (1) the maximum level reached at least 14 days after completion of the initial vaccine regimen but before the additional vaccine dose (labeled “Max Pre”); (2) the last titer measured prior to the additional vaccine dose (“Last Pre”); and (3) the first titer measured after the additional vaccine dose (“First Post”). In analyses, strata of ≥20, 7-<20, 1-<7, and <1 Index were used. The threshold of 20 reflects the semi-quantitative nature of the assay used; the threshold of 7 represents a hypothesized level of protection from severe disease^5^; and the threshold of 1 reflects the manufacturer-recommended lower limit for defining seroresponse.^6^

This study was reviewed and approved by the WCG IRB Work Order 1-1456342-1. Statistical analyses were performed using R v4.0.2.

## Results

A total of 399 patients had received an additional vaccine dose. Only four patients received Ad26.COV2.S/Janssen as the initial vaccine regimen and were therefore excluded from further analysis (**Supplemental Figure 1**). Of the remaining 395 patients, 294 (74%) received a homologous vaccine regimen (**Supplemental Figure 2**). Baseline characteristics are shown in **Supplemental Table 1**.

Among the 395 third dose recipients, 336 (84%) had maximal antibody response ≥7 after the initial series, and 203 (51%) had titer ≥7 just prior to the third dose, measured median 16 [IQR 12-28] before the third dose (**Table 1A**). Following administration of the third dose, 384 (97%) patients had seroresponse ≥7, measured median 33 [IQR 21-40] days later. Among the 37 patients whose maximal response after the initial regimen was only 1 to <7, 36 (97%) had titer ≥7 following the third dose. Among the 22 patients who had titer <1 after the initial regimen, 18 (82%) had an increased response following the third dose, with 14 (64%) developing titers ≥7. Analyses were repeated for subgroups by COVID-19 history (**Tables 1B-C**) and by vaccine type (**Supplemental Table 2**); results were not notably different, though numbers were limited for patients with COVID-19 history.

**Table 1.**
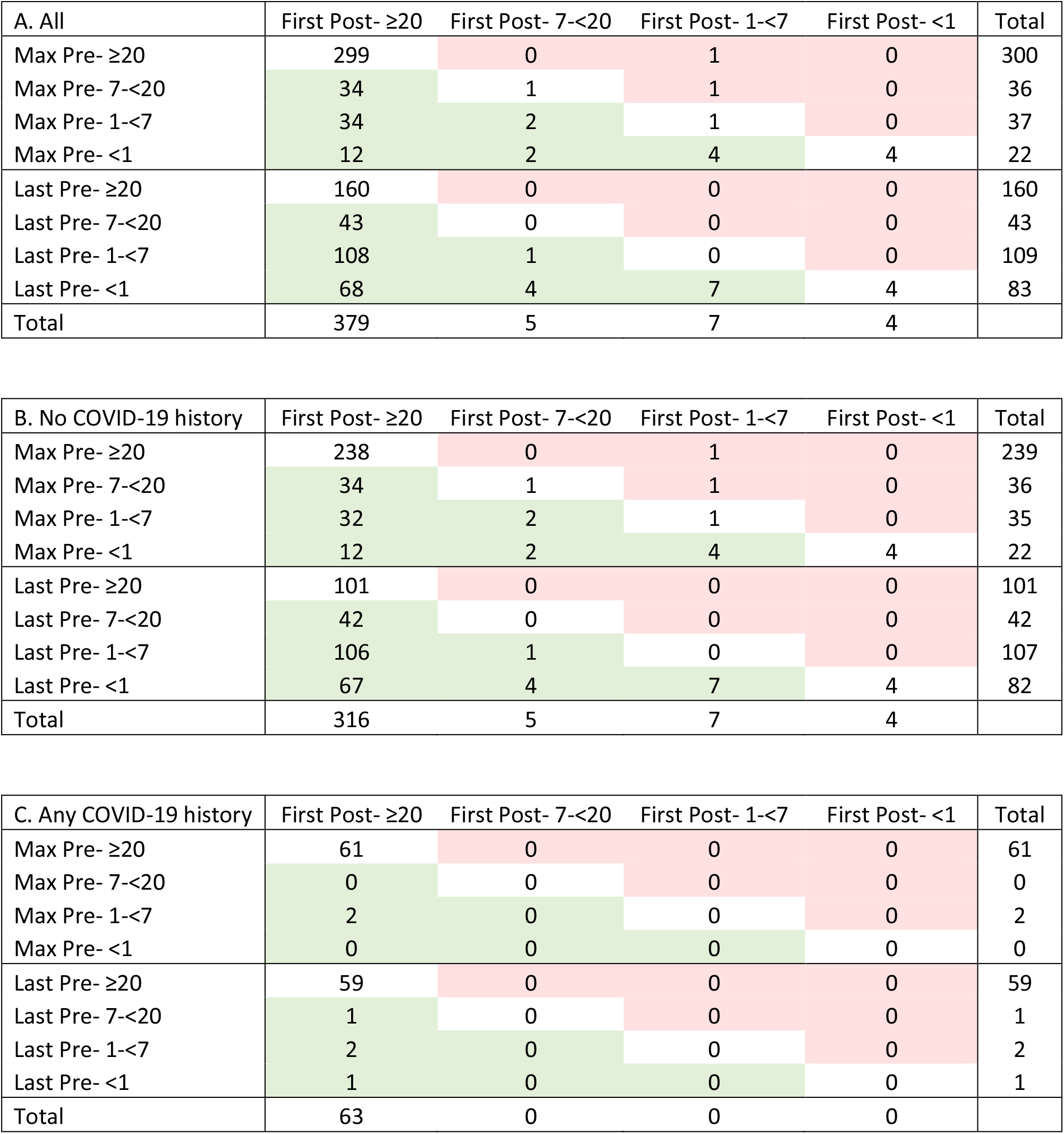
Maximum pre-third dose titer and last pre-third dose titer compared to first post-third dose titer A. All patients B. Patients without history of COVID-19 C. Patients with history of COVID-19 Red cells indicate decrease in stratum from pre- third dose to post- third dose. Green cells indicate increase in stratum from pre- third dose to post- third dose.

## Discussion

Among maintenance dialysis patients, a third dose of mRNA SARS-CoV-2 vaccine is associated with a high prevalence of seroresponse. Previous studies have shown that, while patients receiving maintenance dialysis are able to mount a robust initial immune response, this seroresponse wanes over time.^7^ Studies of the BNT162b2/Pfizer vaccine in France showed that a third dose administered shortly after a second dose was associated with increased seroresponse^8–10^ The current study includes a larger cohort with representation of both mRNA-1273/Moderna and BNT162b2/Pfizer recipients. The data further demonstrate robust seroresponse with titers ≥7 in 97% of recipients of a third vaccine dose, including 97% and 64% of those with suboptimal (<7) or minimal (<1) response, respectively, to an initial mRNA vaccine regimen.

Given ongoing high rates of COVID-19 and rapidly waning immunity after an initial vaccine series, additional or booster doses of SARS-CoV-2 vaccine provided to all maintenance dialysis patients likely will have substantial benefits. Adequate seroimmunity is thought to both reduce breakthrough infections and hospitalizations, and reduce transmission by those who have the infection;^11–13^ all are crucial for patients receiving maintenance dialysis, who are at increased risk for poor outcomes.

This study has limitations, including lack of data on cellular immunity and possible selection bias, both in the enrollment for antibody monitoring and for the administration of additional vaccine dose. We did not correlate antibody levels to breakthrough infections. Importantly, a major strength is that this study includes real-world results on seroresponse to additional doses of SARS-CoV-2 vaccination.

In conclusion, additional SARS-CoV-2 vaccine doses elicit robust seroresponse among patients receiving maintenance dialysis, including among patients whose seroresponse has lapsed, suggesting that additional vaccine doses are critical to maximize and sustain protection of maintenance dialysis patients. In combination with omicron-variant dominance,^14^ we recommend that a third vaccine dose be standard for all maintenance dialysis.

## Data Availability

Data are not available for sharing.

