## Supplemental Tables and Figures for "Seroresponse to third doses of SARS-CoV-2 vaccine among patients receiving maintenance dialysis"

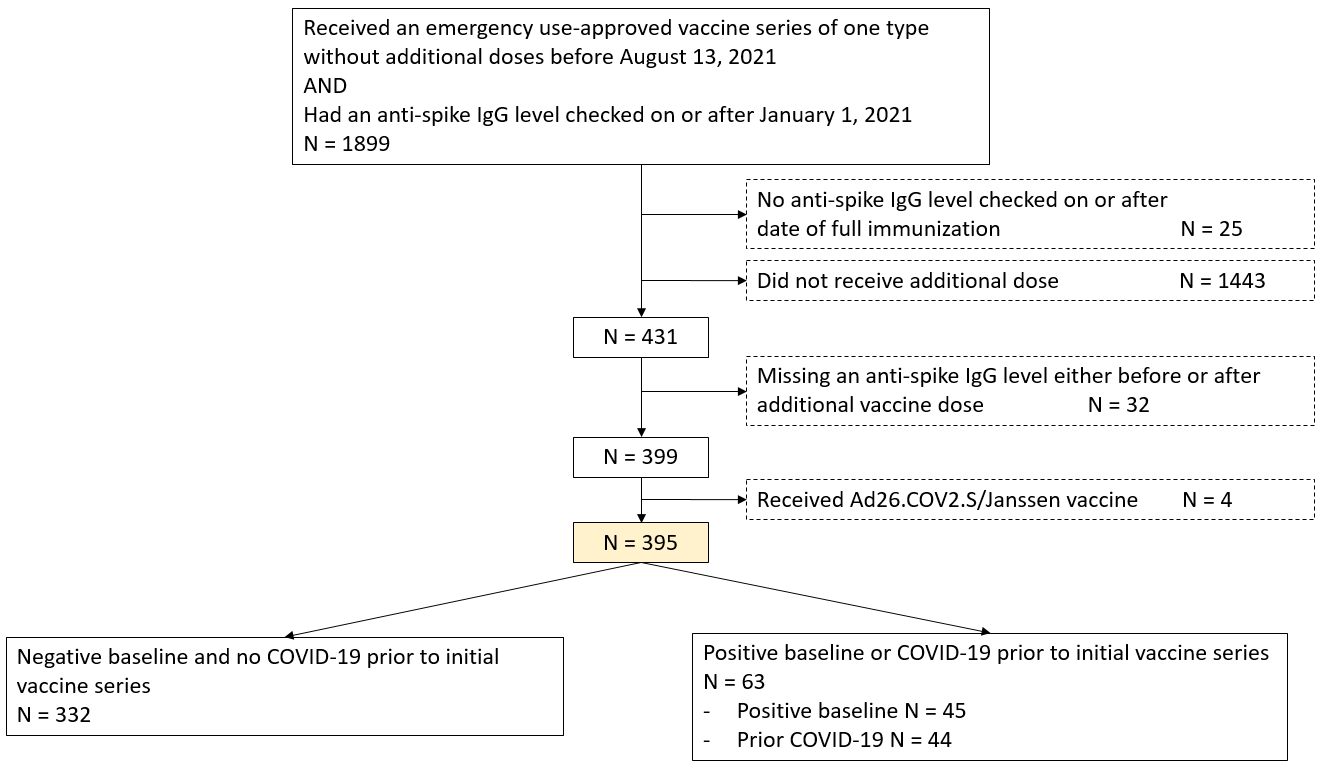

**Supplemental Figure 1. Flow diagram**

Baseline defined as anti-spike IgG titer > 1 Index before or within 10 days after first dose of vaccine (representing likely prior COVID-19 which may or may not have been diagnosed)

Prior COVID-19 defined as positive SARS-CoV-2 test before full immunity (at 14 days after completion of initial vaccine series)

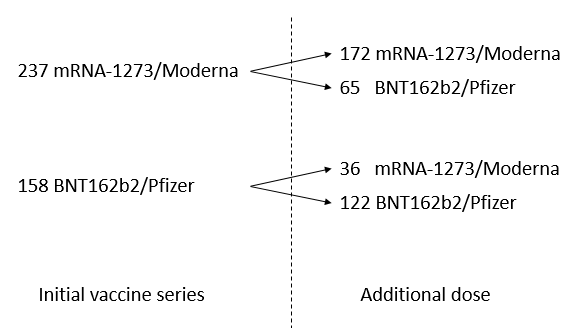

**Supplemental Figure 2. Vaccine regimens of study patients**

**Supplemental Table 1: Baseline characteristics by vaccine regimen**

|  | Overall | MMM^a^ | MMP^a^ | PPM^a^ | PPP^a^ |
| --- | --- | --- | --- | --- | --- |
|  | 395 | 172 | 65 | 36 | 122 |
| Age | 64.1 ± 14.1 | 60.2 ± 13.5 | 68.6 ± 14.6 | 65.5 ± 12.2 | 66.7 ± 13.8 |
| Male sex | 241 (61.0) | 110 (64.0) | 35 (53.8) | 21 (58.3) | 75 (61.5) |
| Race |  |  |  |  |  |
| Native American | 48 (12.2) | 21 (12.2) | 0 (0.0) | 25 (69.4) | 2 (1.6) |
| Asian/Pacific Islander | 28 (7.1) | 3 (1.7) | 2 (3.1) | 0 (0.0) | 23 (18.9) |
| Black | 83 (21.0) | 33 (19.2) | 14 (21.5) | 3 (8.3) | 33 (27.0) |
| Unknown/Other | 39 (9.9) | 13 (7.6) | 16 (24.6) | 1 (2.8) | 9 (7.4) |
| White | 197 (49.9) | 102 (59.3) | 33 (50.8) | 7 (19.4) | 55 (45.1) |
| Hispanic ethnicity | 71 (18.0) | 47 (27.3) | 2 (3.1) | 6 (16.7) | 16 (13.1) |
| Vintage, months | 36 [15, 72] | 38 [16, 69] | 27 [13, 62] | 50 [21, 107] | 32 [14, 68] |
| Body mass index, kg/m^2^ | 28.8 ± 7.4 | 29.2 ± 8.4 | 29.2 ± 6.4 | 29.1 ± 6.7 | 27.8 ± 6.7 |
| Diabetes | 240 (60.8) | 112 (65.1) | 34 (52.3) | 28 (77.8) | 66 (54.1) |
| Long-term care facility | 53 (13.4) | 28 (16.3) | 5 (7.7) | 1 (2.8) | 19 (15.6) |
| Modality |  |  |  |  |  |
| Home hemodialysis | 4 (1.0) | 3 (1.7) | 0 (0.0) | 0 (0.0) | 1 (0.8) |
| In-center hemodialysis | 380 (96.2) | 168 (97.7) | 60 (92.3) | 36 (100.0) | 116 (95.1) |
| Peritoneal dialysis | 11 (2.8) | 1 (0.6) | 5 (7.7) | 0 (0.0) | 5 (4.1) |
| Inadequate dialysis ^b^ | 19 (4.8) | 5 (2.9) | 7 (10.8) | 0 (0.0) | 7 (5.7) |
| Albumin, g/dL | 4 (0.4) | 4 (0.4) | 4 (0.4) | 4 (0.3) | 4 (0.4) |
| HBsAb ≥10 mIU/mL ^c^ | 321 (81.3) | 149 (86.6) | 48 (73.8) | 30 (83.3) | 94 (77.0) |
| History of transplantation | 27 (6.8) | 13 (7.6) | 1 (1.5) | 0 (0.0) | 13 (10.7) |
| Immunodeficiency | 16 (4.1) | 7 (4.1) | 2 (3.1) | 0 (0.0) | 7 (5.7) |
| Immunomodulating medication ^d^ | 56 (14.2) | 22 (12.8) | 11 (16.9) | 1 (2.8) | 22 (18.0) |
| Congestive heart failure | 65 (16.5) | 35 (20.3) | 12 (18.5) | 3 (8.3) | 15 (12.3) |
| Peripheral vascular disease | 34 (8.6) | 20 (11.6) | 1 (1.5) | 4 (11.1) | 9 (7.4) |
| Cerebrovascular disease | 26 (6.6) | 14 (8.1) | 3 (4.6) | 2 (5.6) | 7 (5.7) |
| Chronic obstructive pulmonary disease | 59 (14.9) | 27 (15.7) | 16 (24.6) | 3 (8.3) | 13 (10.7) |
| History of cancer | 43 (10.9) | 16 (9.3) | 6 (9.2) | 0 (0.0) | 21 (17.2) |
| History of COVID-19 | 63 (15.9) | 32 (18.6) | 4 (6.2) | 4 (11.1) | 23 (18.9) |
| Time between initial vaccine regimen and additional dose, days | 169 [148, 196] | 149 [132, 163] | 171 [157, 173] | 204 [185, 219] | 195 [177, 196] |

Vintage and time between initial vaccine regimen and additional dose are reported as median [IQR]. All other data are reported as mean ± standard deviation or %.

Data on baseline patient characteristics were complete.

^a^ The three-letter initials indicate the vaccine regimen, e.g. “MMM” signifies receipt of an initial two-dose mRNA-1273/Moderna vaccine series, followed by a mRNA-1273/Moderna booster

^b^ Inadequate dialysis defined by hemodialysis dose spKt/V<1.2 or peritoneal dialysis dose weekly Kt/V<1.7

^c^ HBsAb ≥10 mIU/mL signifies hepatitis B seroimmunity

^d^ Immunomodulating medications include anti-inflammatory medications, anti-neoplastic agents, corticosteroids, and certain anti-infective medications

| MMM | First Post- ≥20 | First Post- 7-<20 | First Post- 1-<7 | First Post- <1 | Total |  | MMP | First Post- ≥20 | First Post- 7-<20 | First Post- 1-<7 | First Post- <1 | Total |
| --- | --- | --- | --- | --- | --- | --- | --- | --- | --- | --- | --- | --- |
| Max Pre- ≥20 | 142 | 0 | 1 | 0 | 143 |  | Max Pre- ≥20 | 59 | 0 | 0 | 0 | 59 |
| Max Pre- 7-<20 | 8 | 0 | 0 | 0 | 8 |  | Max Pre- 7-<20 | 4 | 0 | 0 | 0 | 4 |
| Max Pre- 1-<7 | 11 | 2 | 1 | 0 | 14 |  | Max Pre- 1-<7 | 1 | 0 | 0 | 0 | 1 |
| Max Pre- <1 | 5 | 0 | 1 | 1 | 7 |  | Max Pre- <1 | 0 | 0 | 0 | 1 | 1 |
| Last Pre- ≥20 | 93 | 0 | 0 | 0 | 93 |  | Last Pre- ≥20 | 30 | 0 | 0 | 0 | 30 |
| Last Pre- 7-<20 | 23 | 0 | 0 | 0 | 23 |  | Last Pre- 7-<20 | 11 | 0 | 0 | 0 | 11 |
| Last Pre- 1-<7 | 30 | 0 | 0 | 0 | 30 |  | Last Pre- 1-<7 | 17 | 0 | 0 | 0 | 17 |
| Last Pre- <1 | 20 | 2 | 3 | 1 | 26 |  | Last Pre- <1 | 6 | 0 | 0 | 1 | 7 |
| Total | 166 | 2 | 3 | 1 |  |  | Total | 64 | 0 | 0 | 1 |  |
| PPP | First Post- ≥20 | First Post- 7-<20 | First Post- 1-<7 | First Post- <1 | Total |  | PPM | First Post- ≥20 | First Post- 7-<20 | First Post- 1-<7 | First Post- <1 | Total |
| Max Pre- ≥20 | 76 | 0 | 0 | 0 | 76 |  | Max Pre- ≥20 | 22 | 0 | 0 | 0 | 22 |
| Max Pre- 7-<20 | 19 | 1 | 1 | 0 | 21 |  | Max Pre- 7-<20 | 3 | 0 | 0 | 0 | 3 |
| Max Pre- 1-<7 | 13 | 0 | 0 | 0 | 13 |  | Max Pre- 1-<7 | 9 | 0 | 0 | 0 | 9 |
| Max Pre- <1 | 6 | 2 | 3 | 1 | 12 |  | Max Pre- <1 | 1 | 0 | 0 | 1 | 2 |
| Last Pre- ≥20 | 30 | 0 | 0 | 0 | 30 |  | Last Pre- ≥20 | 7 | 0 | 0 | 0 | 7 |
| Last Pre- 7-<20 | 8 | 0 | 0 | 0 | 8 |  | Last Pre- 7-<20 | 1 | 0 | 0 | 0 | 1 |
| Last Pre- 1-<7 | 48 | 1 | 0 | 0 | 49 |  | Last Pre- 1-<7 | 13 | 0 | 0 | 0 | 13 |
| Last Pre- <1 | 28 | 2 | 4 | 1 | 35 |  | Last Pre- <1 | 14 | 0 | 0 | 1 | 15 |
| Total | 114 | 3 | 4 | 1 |  |  | Total | 35 | 0 | 0 | 1 |  |

**Supplemental Table 2**. Maximum pre-third dose titer and last pre-third dose titer compared to first post-third dose titer, by vaccines received

The three-letter initials indicate the vaccine regimen, e.g. “MMM” signifies receipt of an initial two-dose mRNA-1273/Moderna vaccine series, followed by a mRNA-1273/Moderna booster.

Red cells indicate decrease in stratum from pre- third dose to post- third dose.

Green cells indicate increase in stratum from pre- third dose to post- third dose.
